# Implicit reconstruction of gating strategies from sample-level flow cytometry annotations

**DOI:** 10.64898/2026.10.20.26365931

**Authors:** Maximilian Buser, Monika Szczepanowski, Anna Laqua, Guranda Chitadze, Claudia Baldus, Monika Brüggemann, Carsten Marr

## Abstract

Flow cytometry is central to clinical hematology, generating extensive archives comprising billions of single-cell profiles linked to expert-derived diagnostic summaries. While these records represent a largely untapped repository of collective expert knowledge, the underlying single-cell annotations required for supervised machine learning are typically not recorded in a standardized, accessible format. We present IMPGAT (implicit gating), a weakly supervised multiple-instance learning framework that reconstructs single-cell gating strategies directly from existing sample-level cell-type frequency distributions. The model utilizes an induced set attention mechanism to map single-cell marker profiles to specific cell populations, requiring neither explicit cell-level labels nor unsupervised clustering during training. We evaluate the framework on acute lymphoblastic leukemia (ALL) and acute myeloid leukemia (AML) cohorts, totaling 711 measurements from 268 individuals and 240 million single cells. In ALL, the model’s quantification of cell-type abundances shows no statistically significant difference from human inter-expert variability. In AML, the framework achieves blast quantification on par with a recently established supervised state-of-the-art model that relies on explicit single-cell annotations for training. Error analysis shows that mismatches between model predictions and expert gating primarily involve expert-labeled non-cellular events, reflecting the non-incorporation of any explicit physical prior knowledge (such as the geometric relationships traditionally used to identify doublets and debris in scatter cytograms) in model training. Conversely, model certainty correlates with phenotypic borderline cases where manual bisection is inherently difficult. Our approach enables the systematic utilization of retrospective clinical records for the development of automated, standardized gating logic and is applicable to broader high-throughput contexts, such as large-scale immune monitoring or phenotypic drug screening.

## Main

Flow cytometry is a fundamental tool in hematology research [1] and diagnostics [2, 3], essential for the in-depth phenotypic characterization of (pathological) cell populations. As a result, extensive clinical archives exist containing raw flow cytometry measurements alongside corresponding diagnostic reports. While these reports provide diagnostically relevant sample-level statistics, like cell-type frequencies, explicit single-cell annotations are rarely recorded in standardized or accessible formats, complicating the re-use of data for model training [4].

Over the last decade, machine learning (ML) approaches – encompassing both supervised and unsupervised paradigms – have emerged to support [5–7] or even fully automate [8, 9] flow cytometry analysis. For instance, deep representation frameworks such as cytoVI [10] – adapted from single-cell transcriptomics architectures [11] – have significantly advanced the mapping, batch correction, and analysis of flow cytometry data featuring phenotypic continuums. However, fully unsupervised, clustering-based methods often struggle to align with established biological gating hierarchies [12]. While they have proven useful for expert-guided interactive data analysis [13], the requirement for manual post-processing and interpretation limits their utility for routine hematological assessment, such as the quantification and characterization of leukemic blasts in acute myeloid leukemia (AML) or acute lymphoblastic leukemia (ALL).

B-cell precursor ALL exemplifies these diagnostic challenges; here, the precise identification of CD20-positive versus CD20-negative leukemic blast fractions is a critical determinant of rituximab eligibility [14]. Recent findings from the GMALL 08/2013 trial (NCT02881086) demonstrate that CD20 expression on leukemic blasts is highly dynamic, frequently shifting across treatment phases and body compartments (e.g., bone marrow versus peripheral blood) [15, 16]. This phenotypic het-erogeneity precludes the use of static thresholds, leading to inter-expert variability in manual gating, and underscoring the need for data-driven approaches to faithfully resolve phenotypic populations [17, 18]. In AML, supervised approaches, such as those utilizing Gaussian mixture models [19], have reached high concordance with manual expert gating in the (semi)-automated identification of leukemic blasts [20].

However, these models require labor-intensive, expert-provided single-cell labels for training, annotations that are difficult to scale and not available for most large clinical cohorts or retrospective archives. To address these limitations, we propose the IMPGAT (implicit gating) model, a weakly supervised framework designed to utilize retrospective clinical archives that contain sample-level cell-type frequencies but lack explicit single-cell annotations. While previous weakly supervised frame-works like CellCnn [21] or CytoDX [22] successfully exploited sample-level metadata to flag disease-associated cellular subsets, IMPGAT introduces a paradigm to back-calculate full single-cell gating strategies. The core hypothesis is that the variance in cell-type frequencies across a sufficiently large cohort provides an implicit supervision signal, enabling the reconstruction of the underlying single-cell gating logic from sample-level labels only. IMPGAT is based on a multiple-instance learning (MIL) architecture [23–25] with a feature extractor utilizing an induced set attention block (ISAB) [26] to maintain the permutation invariance inherent to flow cytometry data. This design treats each flow cytometry measurement of arbitrary length as an unordered set of single cells – reflecting the fact that the recording sequence is purely stochastic – while simultaneously capturing the inter-cell context helping to resolve phenotypic populations relative to the global sample distribution.

## Results

### Reconstruction of expert-gating logic via implicit supervision

To bypass the need for explicit single-cell annotations during training, we developed an implicit gating model. IMPGAT reconstructs single-cell gating logic from raw flow cytometry measurement data and aggregate sample-level labels (Fig. 1 A, B, C). We demonstrate and validate the model’s performance on two distinct cohorts: a B-cell precursor ALL dataset from the GMALL 08/2013 (NCT-02881086) trial (226 measurements from 129 individuals with an average of 7.0 *×* 10^5^ cells per measurement) [15, 16] and a previously published AML dataset (485 measurements from 139 individuals with an average of 1.7 *×* 10^5^ cells per measurement) [19]. ALL samples were measured under standardized EuroFlow instrument settings using an 8-color panel (CD10, CD19, CD20, CD34, CD38, CD45, CD66c+CD123, and CD81) [27–29], enabling the resolution of mutually exclusive cell populations, specifically, CD20+ and CD20-blasts, mature B cells, other lymphocytes, and other nucleated cells, as well as debris and doublets. For the AML dataset, our analysis focuses on the five shared backbone markers (CD13, CD34, CD45, CD117, and HLA-DR) available across all tubes of the underlying multi-tube panel measurements. Using these backbone markers, we resolve three primary populations: leukemic blasts, non-blast white blood cells (WBC), and other events – consistent with the original expert annotations [19].

**Figure 1:**
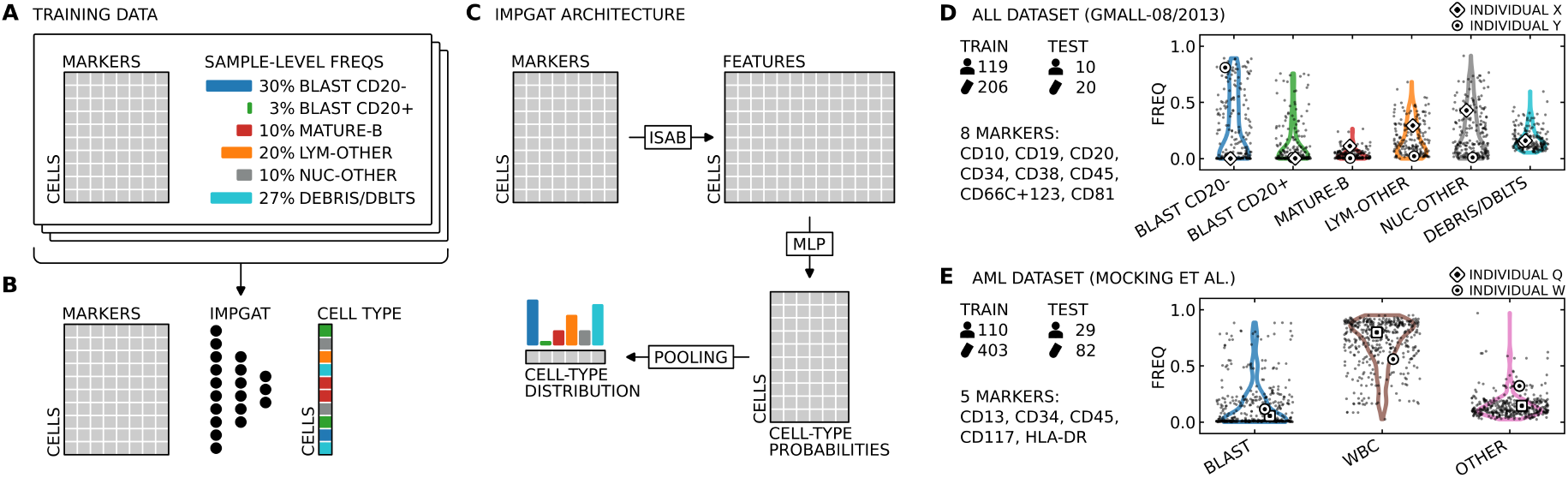
Implicit reconstruction of gating strategies from weakly annotated flow cytometry data. **A**. The IMPGAT model is trained on raw flow cytometry measurements (cells *×* marker expression) where only the aggregate sample-level cell-type frequencies are known, but no single-cell annotations. **B**. The model processes marker expression profiles to predict single-cell labels, i.e., the cell type. **C**. The model utilizes an induced set attention block (ISAB) for permutation-invariant feature extraction. The ISAB realizes a two-stage attention mechanism where a set of learnable inducing points first attends to the single-cell data to capture global sample-level context; second, the cells attend back to these inducing points. This enables the model to account for inter-cell relationships while dealing efficiently with millions of data points. The resulting single-cell features are processed individually by a multi-layer perceptron (MLP) head with a final softmax activation to produce cell-specific cell-type probabilities. Pooling of these single-cell predictions yields the sample-level frequencies that match the labels observed during training. The approach is exemplified for two datasets: **D**. An ALL dataset comprising 226 initial treatment-naïve and post-prophase samples from 129 patients in the GMALL 08/2013 trial (NCT02881086). We use 206 measurements from 119 individuals for training and 20 measurements from 10 individuals for testing. ALL samples were measured using a standardized EuroFlow panel comprising eight markers: CD10, CD19, CD20, CD34, CD38, CD45, CD66c+CD123, and CD81. The training data are annotated with sample-level frequencies for mutually exclusive cell types, including CD20 negative and positive blasts (BLAST CD20-/+), mature B cells (MATURE-B), other lymphocytes (LYM-OTHER), and other nucleated cells (NUC-OTHER), as well as debris and doublets (DEBRIS/DBLTS). Cell-type frequency distributions across the training set are visualized as violin plots overlaid with individual measurement data points. Two representative samples from two individuals are highlighted (square and circle markers) to illustrate the inter-sample heterogeneity in cell-type composition. For all ALL samples, sample-level cell-type frequencies from routine expert gating are available and used for training. For the test set, we further generated single-cell annotations from a second expert gating. **E**. An AML dataset [19] with derived mutually exclusive labels for blasts (BLAST), white blood cells excluding blasts (WBC), and other cells (OTHER). The dataset comprises 403 measurements from 110 individuals for training and 82 measurements from 29 individuals for testing, following the train-test split from the original publication. Measurements in the original study were acquired using a four-tube panel; in this work, we restrict the analysis to the five shared backbone markers (CD13, CD34, CD45, CD117, HLA-DR) that are available across all tubes. The training set includes 18 non-AML control samples.

As the IMPGAT model receives no explicit single-cell labels during training, it relies on the natural variance in cell-type composition across large training data cohorts to derive an implicit supervision signal (Fig. 1 D, E). By optimizing the fit between predicted and observed sample-level cell-type frequencies, the framework learns to map high-dimensional single-cell marker profiles to specific phenotypic populations.

### IMPGAT achieves expert-level performance in ALL analytics

We first assessed whether IMPGAT could faithfully reconstruct established gating hierarchies for B-cell precursor ALL samples. We trained the model on 206 measurements from 119 individuals in a five-fold cross-validation manner (Methods) and evaluated its performance on a hold-out test set spanning 20 measurements from 10 individuals. Qualitative comparison of representative cytograms demonstrates that IMPGAT-derived gating patterns closely mirror manual expert annotations (Fig. 2 A). Quantitatively, the model achieved high classification performance across the test set measurements, with a balanced accuracy of 88.0% *±* 4.7%, and weighted and unweighted F1-scores of 90.4% *±* 6.3% and 84.4% *±* 6.7%, respectively (mean *±* standard deviation).

**Figure 2:**
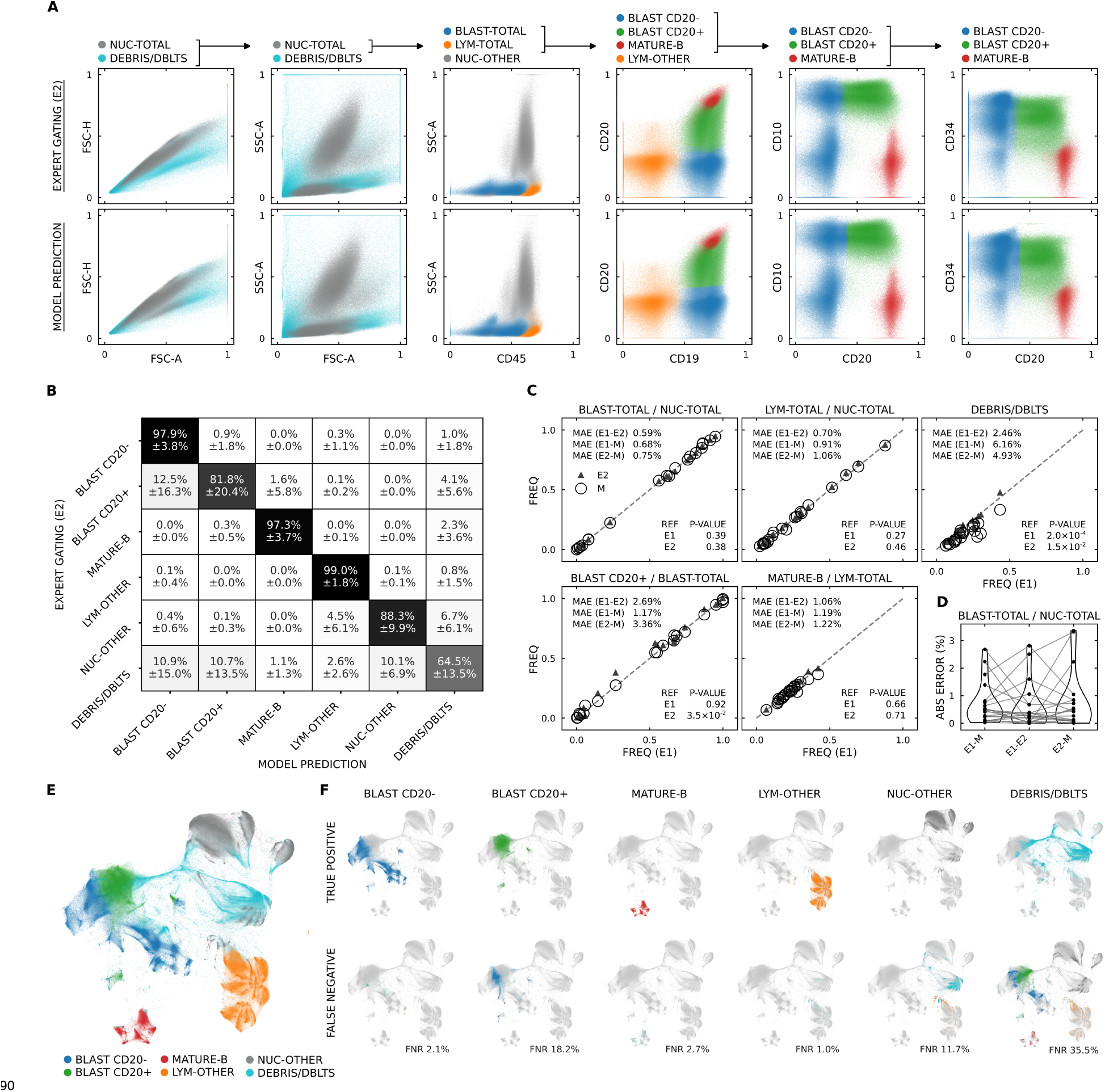
Gating patterns implicitly learned from sample-level labels mirror single-cell expert annotations in ALL. **A**. Expert gating (E2, top row) and model predictions (bottom row) largely coincide in representative cytogram views. Scatterplots display an equisampled aggregate of single cells across all test samples. In addition to the mutually exclusive labels (BLAST CD20-, BLAST CD20+, MATURE-B, LYM-OTHER, NUC-OTHER, and DEBRIS/DBLTS), NUC-TOTAL refers to all labels excluding DEBRIS/DBLTS; LYM-TOTAL accounts for MATURE-B and LYM-OTHER; and BLAST-TOTAL represents BLAST CD20- and BLAST CD20+. **B**. Row-normalized confusion matrix for model predictions versus expert (E2) annotations on the ALL test set. Entries represent mean and standard deviation across test samples. The lower performance observed for DEBRIS/DBLTS reflects the inherent difficulty in establishing manual gating thresholds for non-cellular events. **C**. Regression plots of phenotypic fractions confirm high concordance between model predictions (M) against expert gating results (E1 and E2) on the ALL test set. Insets report mean absolute errors (MAE) alongside p-values from one-sided Wilcoxon signed-rank tests evaluating whether model error significantly exceeds inter-expert variability, benchmarking model error against inter-expert variability using E1 and E2 as alternating references. Except for debris and doublets, the model’s deviations show no significant difference from human inter-expert variability (all *P ≥* 0.035; exceeding *α =* 0.025, which corresponds to a 5% significance threshold Bonferroni-corrected for two tests). **D**. Violin plots of absolute errors for the BLAST-TOTAL/NUC-TOTAL fraction show comparable model deviations against each expert (E1-M, E2-M) next to the baseline inter-expert variability (E1-E2). Solid circles represent individual measurements in the test set; lines connecting them represent corresponding measurements, visualizing the variance evaluated by the error-comparison tests. **E**. Two-dimensional UMAP embedding of single-cells colored by expert labels (E2). **F**. Error analysis of single-cell model predictions with respect to expert labels (E2) projected onto the same UMAP embedding. The top row resolves true positive predictions, while the bottom row shows false negatives; colors in the bottom row indicate the original expert-assigned labels for the misclassified cells; the annotated false negative rate (FNR) accounts for the proportion of expert-annotated cells missed by the model for each label.

The corresponding confusion matrix shows high classification fidelity for all cell types (e.g., a sensitivity of 97.9% for CD20-blasts, and 81.8% for CD20+ blasts), though a notable mismatch was observed for expert-labeled debris and doublets with a false negative rate of 35.5% (Fig. 2 B). This discrepancy highlights an inherent divergence between expert gating – which typically relies on the geometric bisection of scatter cytograms to exclude non-cellular events – and the model’s decision boundaries, which are implicitly learned from correlating sample-level cell-type frequencies with high-dimensional marker distributions without explicit physical prior knowledge.

To evaluate the accuracy of population quantification, we analyzed four diagnostically relevant phenotypic fractions: total blasts/nucleated cells, total lymphocytes/nucleated cells, CD20+ blasts/ total blasts, and mature B cells/total lymphocytes. Using the measurement from two independent expert annotations (E1 and E2), we found high correlations of cell type fractions for both experts and the model predictions, with mean absolute errors (MAE) below 3.5% for diagnostically relevant cell types (Fig. 2 C). Only for the classification of debris and doublets, higher errors occur between the model and the experts (MAE of 6.16% and 4.93%), but also between the experts (MAE of 2.46%). We performed one-sided Wilcoxon signed-rank tests to assess whether model-to-expert deviations remained within the bounds of inter-expert variability. For each of the four diagnostically relevant fractions, the absolute errors showed no statistically significant difference compared to the variability between two independent human expert annotations according to a 5% significance threshold Bonferroni-corrected for two tests (*P ≥* 0.38 for total blasts/nucleated cells, *P ≥* 0.27 for total lymphocytes/nucleated cells, *P ≥* 0.035 for CD20+ blasts/total blasts, and *P ≥* 0.066 for mature B cells/total lymphocytes all exceeding *α =* 0.025) (Fig. 2 D). Conversely, model deviations were significantly higher than inter-expert variability for the classification of debris and doublets (*P =* 2.0 *×* 10^*−*4^ and *P =* 0.015 for the respective expert references), identifying this non-cellular category as the primary source of divergence from human expert-derived gating. UMAP embedding of all test cases shows a complex pattern with distinct clusters (Fig. 2 E) and confirms the ability of the model to identify cell populations (Fig. 2 F, top), with false negatives largely restricted to the interface of CD20+ versus CD20-blasts, and the above-mentioned debris and doublet category (Fig. 2 F, bottom).

### Error analysis and model uncertainty

To elucidate limitations of the IMPGAT model, we performed a detailed error analysis of the ALL test sample where the model performed worst with a balanced accuracy score of 78.5% (Fig. 3 A). The confusion matrix of the sample shows a pronounced confusion of CD20-blasts with CD20+ blasts and non-cellular events (Fig. 3 B), while mature B cells, other lymphocytes, and other nucleated cells are correctly resolved. Interestingly, the model certainty correlates with errors, showing the highest uncertainty where mismatches occur (Fig. 3 C, right). Zooming in, these mismatches primarily stem from events positioned to the right of the diagonal singlet axis in forward scatter height versus area cytograms (Fig. 3 D, 1st and 2nd row), coming from doublet events. While these events were expert-gated as doublets, the model assigned them primarily to the CD20-blast population based on learned marker correlations. Apart from the missed doublet population, we found that the highest levels of model uncertainty coincided with interface regions between neighboring cell populations and phenotypic borderline cases. In these regions, manual bisection of different phenotypes is inherently difficult for continuous density gradients that lack discrete, bimodal partitions (Fig. 3 D, 3rd, 4th, and 5th row). This suggests that the IMPGAT approach helps quantify the ambiguity in gradual phenotypic distributions without clear cluster-based partitions. For the ALL test sample where the model performed best with a balanced accuracy score of 96.0%, blast populations as well as debris and doublets are faithfully resolved (Supplemental Fig. 5).

**Figure 3:**
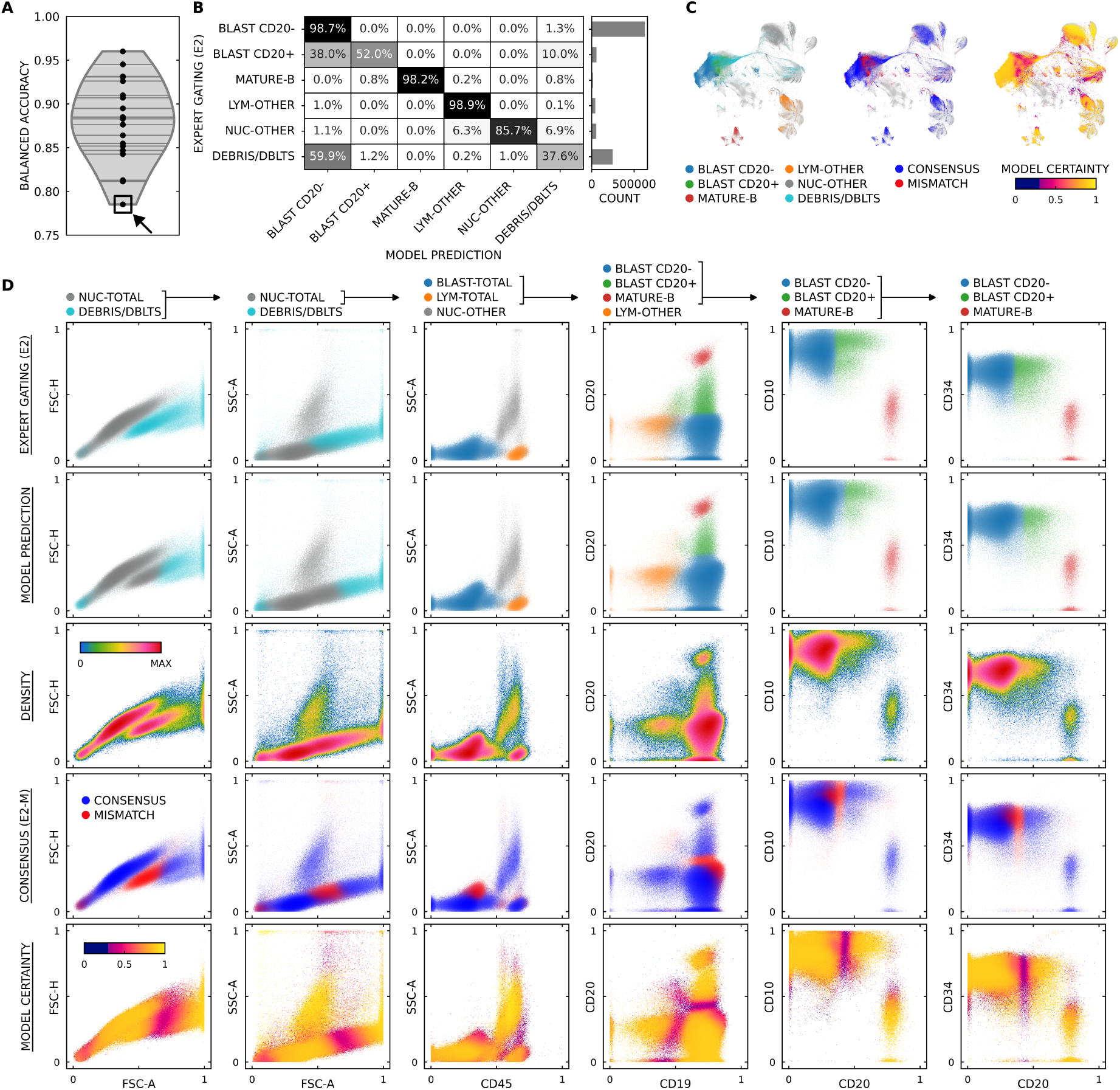
Detailed error analysis of the worst-prediction ALL test sample identifies the classification of non-cellular events as the primary source of model-expert mismatch. **A**. The worst-prediction test sample (square marker), defined by the lowest balanced accuracy, is selected for a detailed error analysis. It is a treatment-naïve bone marrow sample with a high blast burden (60.2% BLAST CD20-, 5.4% BLAST-CD20+). **B**. The sample’s confusion matrix reveals two notable discrepancies: the misclassification of expert-labeled debris and doublet populations as CD20-blasts and a shift between CD20+ and CD20-blast populations. **C**. Projection onto the global test set UMAP (light grey background, compare Fig. 2 E, F) resolves expert-labeled cell types, model-expert consensus, and model certainty. **D**. Representative cytogram views of the worst-prediction sample, organized by population-specific marker and scatter channel projections. Rows resolve expert gating (row 1), model predictions (row 2), pseudo-density (row 3), model-expert consensus (row 4), and model certainty (row 5), with row 1 and row 2 displaying cells according to the respective expert or model labels, and row 3, 4, and 5 displaying the union of these events. The comparison reveals that the primary source of disagreement involves events positioned to the right of the diagonal singlet axis in the FSC-H versus FSC-A projection. While these events are manually gated as debris and doublets, the model – operating without any prior physical knowledge concerning singlet, doublet, and debris scatter characteristics – assigns them to the CD20-blast population based on learned sample-level correlations. The discrepancies between CD20- and CD20+ blast populations occur primarily at the boundary regions of continuous phenotypic distributions, underscoring the inherent difficulty of establishing a discrete manual cutoff in these cases. The single-cell model certainty closely follows expert-model consensus, with the highest uncertainty being observed at interface regions between neighboring cell populations, indicating that model uncertainty serves as a proxy for manual classification difficulty.

### Benchmarking against supervised state-of-the-art in AML blast identification

To assess the generalizability of our framework, we evaluated IMPGAT on an independent AML dataset (Fig. 1 E). Despite the lack of explicit single-cell labels for training purposes, the model faithfully recovered distinct cellular populations (Fig. 4 A), yielding blast and non-blast white blood cell counts that were highly concordant with the original expert annotations (Spearman rank correlation coefficients *r ≥* 0.95; Fig. 4 B). Critically, IMPGAT’s blast quantification performance was on par with a supervised state-of-the-art Gaussian mixture model-based approach [19] (Fig. 4 C).

**Figure 4:**
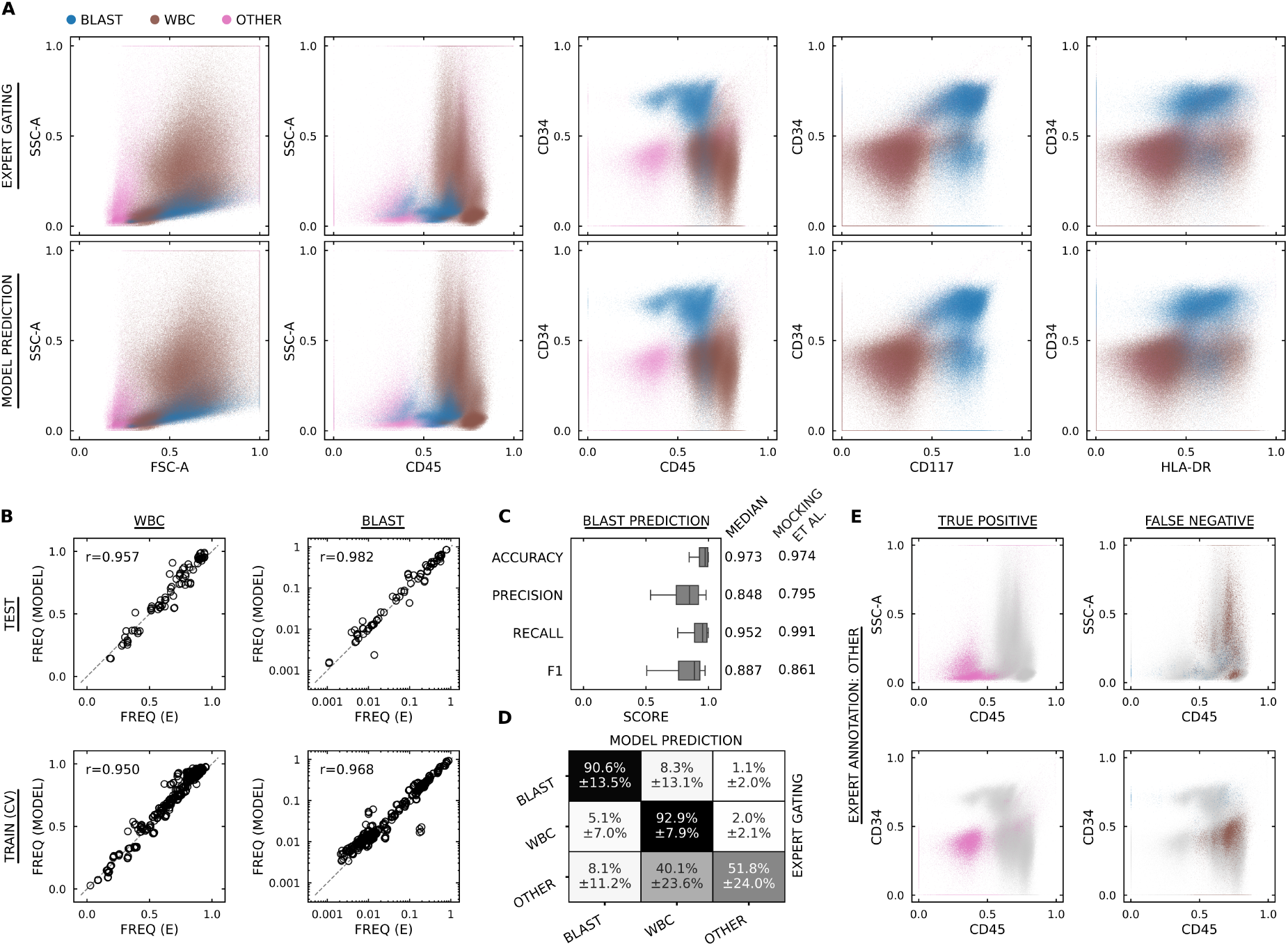
Gating patterns learned from sample-level labels recover expert single-cell populations in AML data. **A**. Comparison of expert gating (top row) and model predictions (bottom row) in characteristic cytogram views. Scatterplots display an equisampled aggregate of single-cell events across all test measurements. **B**. Regression analysis of WBC and BLAST frequencies on the test set (top row) and across train cross-validation folds (bottom row) yield high Spearman rank correlation coefficients *r ≥* 0.95. **C**. Benchmarking of BLAST prediction performance shows that our IMPGAT model, trained using only weak sample-level annotations, achieves results on par with those reported by Mocking et al. [19], which utilized explicit single-cell labels for training a Gaussian mixture model-based blast classifier. **D**. Row-normalized confusion matrix for model predictions on the AML test set. The highest discrepancy is observed in the OTHER category (48.2% false negative rate), which encompasses non-target events including debris. **E**. Representative cytogram views of all test-data events expert-labeled as OTHER, resolving true positive and false negative predictions. Error patterns suggest that a proportion of events labeled as OTHER by experts exhibit scatter and marker profiles characteristic of blast and non-blast white blood cell populations.

**Figure 5:**
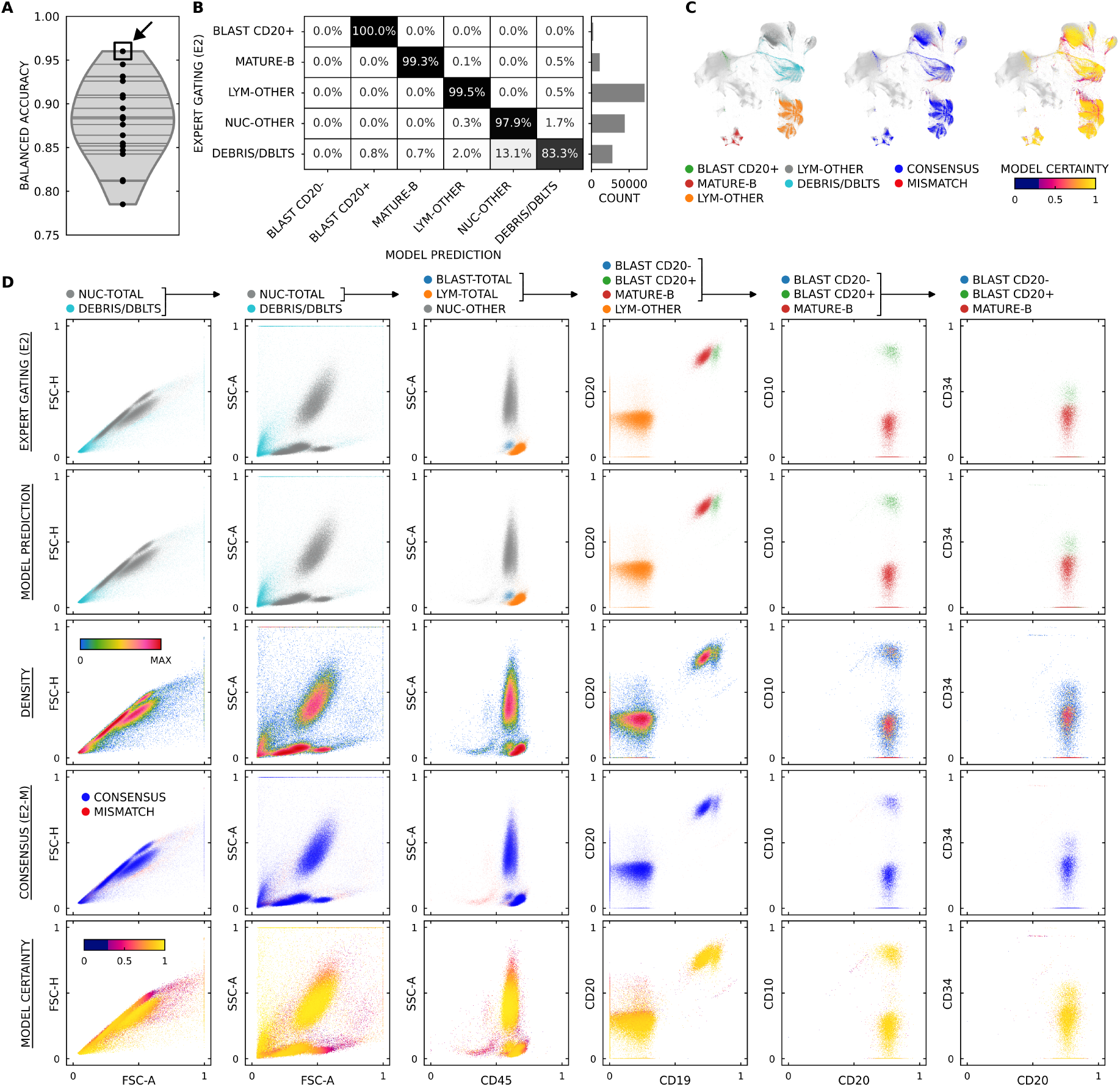
Detailed analysis of the best-prediction ALL test sample. In analogy to Fig. 3, this supplementary figure illustrates the model behavior for the test measurement with the highest balanced accuracy. Note that the BLAST CD20-row is absent from the confusion matrix (B) as this population was not identified in the expert gating of this specific measurement. The underlying measurement is from a post-prophase peripheral blood sample (Methods).

Consistent with our findings in the ALL cohort, analysis of the AML confusion matrix revealed that the highest discrepancy between expert gating and model predictions was observed for the OTHER category (a heterogeneous class encompassing also non-cellular events) which exhibited a false negative rate of 48.2% (Fig. 4 D). Visual inspection of these expert-labeled OTHER events in standard cytogram views suggests that a considerable proportion exhibits scatter and marker profiles characteristic of blast and non-blast white blood cells (Fig. 4 E). Overall, these findings demonstrate that the variance in cell-type frequencies in large-scale cohorts provides an implicit supervision signal that is sufficient to achieve quantification performance on par with models requiring explicit single-cell annotations for training.

## Discussion

Clinical flow cytometry archives represent an expansive resource for the development of automated diagnostics and predictive models, as they contain raw single-cell measurements paired with diagnostically relevant cell-type frequencies. By correlating these sample-level labels with high-dimensional marker profiles across large cohorts, our IMPGAT framework derives an implicit supervision signal to reconstruct single-cell gating strategies. This approach enables the systematic utilization of retrospective clinical records for machine learning, circumventing the requirement for labor-intensive manual single-cell annotations that can otherwise constrain the development of flow cytometry-based models, in the domain of hematological diagnostics and beyond.

The performance of IMPGAT was validated across two distinct clinical contexts and marker panels. In B-cell precursor ALL, phenotypic ambiguity (e.g., concerning the CD20 expression of leukemic blasts) often complicates manual analysis and interpretation; here, IMPGAT’s quantification of phenotypic fractions that are diagnostically relevant was statistically non-different from human inter-expert variability. The framework’s generalizability was further demonstrated in an AML cohort, where the model achieved parity in blast quantification with a supervised state-of-the-art Gaussian mixture model-based approach. Intriguingly, error analysis of the AML data revealed that some events expert-labeled as OTHER exhibited immunophenotypic profiles characteristic of white blood cells and blasts. Overall, these findings indicate that the inter-sample variance in cell-type composition inherent to clinical cohorts provides a sufficient signal to accurately delineate cellular populations across different diagnostic contexts and panels. By learning from large retrospective cohorts, IMPGAT captures a consensus representation of expert practice rather than reproducing the decisions of a single annotator. This provides a principled approach to reducing subjective variability and promoting inter-laboratory standardization.

A key advantage of our purely data-driven approach is its ability to quantify the ambiguity inherent in complex phenotypic distributions. Biological populations frequently exist as phenotypic continua rather than discrete, well-separated clusters [30]. While traditional manual gating necessitates an inherently subjective bisection of these continuous density gradients, IMPGAT’s uncertainty metric directly correlates with these borderline cases. By providing a probabilistic measure of classification difficulty, the framework can serve as an automated quality-control mechanism, identifying samples where manual bisection is likely to be inconsistent and where additional expert review may be in order.

Detailed error analysis identifies the classification of non-cellular events as the primary source of divergence from human expert gating. This mismatch reflects the framework’s reliance on high-dimensional correlations without incorporating the explicit physical prior knowledge – such as scatter-based geometric relationships – traditionally used to resolve nucleated single cells. While omitting these constraints ensures the flexibility of the core IMPGAT methodology, clinical-grade implementations could incorporate additional supervised 2D-projection gating [31, 32] for site-specific debris and doublet removal. Nevertheless, this divergence highlights that data-driven thresholds based on multi-parametric profiles may capture immunophenotypically relevant events that are traditionally excluded by rigid, scatter-based exclusion criteria.

As an implicit regression framework, the performance of IMPGAT is intrinsically linked to the heterogeneity of the training cohort. The model requires sufficient variation in cell-type frequencies across samples to derive a meaningful supervision signal. For instance, if a training cohort lacked phenotypic diversity – such as a dataset where every sample contained near-identical blast fractions – the learning signal would be insufficient to resolve faithful population boundaries. In this paradigm, sufficient big data is defined not only by the absolute number of samples and cells, but also by the phenotypic variance within the training cohort.

There are no inherent architectural barriers preventing the application of IMPGAT to even higher-dimensional data, such as spectral flow [33] or mass cytometry [34]. As these technologies transition from research settings into clinical routine, the ability to derive standardized gating logic from existing diagnostic reports will become increasingly vital. By distilling a consensus gating logic from the aggregate expertise stored in large-scale clinical archives, IMPGAT mitigates the subjectivity inherent in manual analysis. While the utility of the framework is demonstrated here for hematological disorders, the underlying methodology is equally applicable to other high-throughput contexts where sample-level metadata is available, such as large-scale immune monitoring [35] or ex vivo drug screening [36, 37].

## Methods

### Data collection

The B-cell precursor ALL dataset (Fig. 1 D) was derived from a subset of individuals enrolled in the GMALL 08/2013 trial (NCT02881086), a risk-adapted, MRD-stratified therapy-optimization trial for adults (aged 18–55 years) with newly diagnosed ALL. We utilized 226 flow cytometry measurements from 129 individuals, encompassing both treatment-naïve and early treatment stages. Specifically, the cohort included baseline bone marrow and blood samples collected at day 0 (before therapy), as well as blood samples collected at day 6 following a cyclophosphamide- and dexamethasone-containing prophase before induction therapy [16]. All samples were processed and measured with a BD FACSLyric cytometer according to standardized 8-color EuroFlow instrument settings [27–29] using a panel spanning CD10, CD19, CD20, CD34, CD38, CD45, CD66c+CD123, and CD81 antibodies. For the development and evaluation of the IMPGAT framework, the dataset was split at the individual level to prevent information leakage between training and testing. The training set comprised 206 measurements from 119 individuals, while the independent test set consisted of 20 measurements from 10 individuals. All participants provided written informed consent, and the study was approved by the institutional ethics committee in accordance with the Declaration of Helsinki.

For the AML dataset (Fig. 1 E), we used a previously published cohort [19], consisting of 485 measurements from 139 individuals. To ensure comparability of results, we maintained the original training and independent test set split as defined in the original study. From the original expert gating annotations, we distilled three mutually exclusive cell-type labels for model training and evaluation: (leukemic) blasts, non-blast WBC, and a category for other events (encompassing non-cellular events like debris and doublets). For all measurements, the analysis was restricted to the five shared backbone markers (CD13, CD34, CD45, CD117, and HLA-DR) available across all tubes of the underlying multi-tube panel.

### Expert gating of ALL data

To establish a ground truth for both model training and performance evaluation, we utilized two distinct tiers of expert annotations for the ALL cohort: retrospective sample-level labels for the training and the test set (E1) and independent, event-level annotations for the test set (E2).

For the weak supervision of the IMPGAT model, we used sample-level cell-type frequencies derived from manual, per-sample gating of each individual measurement performed by a senior expert (E1) in diagnostic flow cytometry [16]. From the expert’s hierarchical gating, we distilled six mutually exclusive cell-type labels for model training: CD20+ blasts, CD20-blasts, mature B cells, other lymphocytes, and other nucleated cells. A sixth category, debris and doublets, was used to capture all remaining events not classified into the specific cellular populations. To evaluate the model’s performance at the single-cell level, a second set of independent event-level annotations was generated for the measurements in the test set. A second expert (E2), blinded to both the E1 results and the model’s predictions, performed manual gating for each measurement file individually. This provided event-level labels for the test data, which contained an average and median of 7.1 *×* 10^5^ and 4.8 *×* 10^5^ cells per measurement file, respectively. These dual test set annotations allowed us to assess whether the model’s implicitly derived gating logic remained within the bounds of standard inter-expert variability for sample-level metrics and to perform the detailed single-cell error analysis presented in this study.

### Data preprocessing for model training and analysis

Raw flow cytometry data in the FCS 3.1 standard [38] were processed to generate a normalized input representation for the IMPGAT model and data analysis. Initial preprocessing began with the application of the hardware-derived compensation matrix provided in the FCS file headers to correct for fluorescence spillover; no custom post-compensation was applied. Subsequently, each measurement was independently normalized to the [0, 1] range using the maximum range parameters stored in its respective FCS metadata. Fluorescence marker channels underwent a consecutive logarithmic transformation with a small offset (1.0*×*10^*−*5^), while scatter channels (forward and side scatter height and area) were maintained in linear space. Following the logarithmic transformation, a final linear rescaling to the [0, 1] range was applied. This per-sample transformation of marker and scatter intensities ensured that no information from the training cohort influenced the test set representation. This transformed representation of marker and scatter intensities was used consistently throughout this study for both the machine learning analyses and the generation of all cytogram visualizations.

### IMPGAT model architecture

The IMPGAT model is implemented as a multiple-instance learning regression framework designed to process flow cytometry measurements as unordered sets of single-cell vectors (Fig. 1). To maintain the permutation invariance and capture per-sample inter-cell context, the architecture utilizes a set transformer-based feature extractor [26]. Each measurement is treated as a bag of single-cell instances, where each instance is a vector of normalized marker and scatter intensities.

The IMPGAT model architecture consists of three stages: First, an *N × D*-dimensional flow cytometry measurement input matrix (*N* cells, *D* markers and forward and side scatter channels) is projected into a higher dimensional (*N×*128) embedding space to provide sufficient capacity for capturing non-linear relationships (Fig. 1 C). This is followed by an induced set attention block ISAB [26], which employs a set of *M =* 32 learnable inducing points to aggregate global measurement context across the entire cell population. This mechanism allows the model to resolve phenotypic populations relative to the global sample distribution while maintaining a computational complexity of *O*(*N × M* ), circumventing the *O*(*N* ^2^) complexity of standard self-attention, which is computationally prohibitive for typical flow cytometry measurement data comprising 10^5^ or more events. Second, the 128-dimensional context-aware single-cell embeddings are passed through a multi-layer perceptron (MLP) head consisting of two hidden layers (128 and 64 units) with ReLU activations. A final linear layer with a softmax activation generates a probability distribution for each cell across the predefined cell-type categories. Third, to derive sample-level statistics, mean pooling is applied to the individual cell-type probabilities, yielding a predicted frequency vector for the entire measurement.

The model is trained using an implicit supervision signal by minimizing the Kullback-Leibler divergence between the predicted and ground-truth sample-level cell-type frequencies.

### IMPGAT model training

We employed a five-fold cross-validation strategy on the training cohort, with a split at the individual level. For each of the five folds, an ensemble of models was optimized using a nested validation strategy.

For each split, the model was optimized using an Adam optimizer with a learning rate of 1.0 *×* 10^*−*3^. Training was conducted for a maximum of 500 epochs, with the best model state selected based on the minimum validation loss, defined as the Kullback-Leibler divergence of ground-truth and predicted cell-type frequency distributions. To balance the training bags while maintaining representative cellular distributions, we sampled 20,000 cells per measurement per training iteration. The models were trained with a batch size of 8 flow cytometry measurements.

For the final evaluation on the hold-out test set, we utilized a pooled ensemble approach. Predictions from the optimized model instances across all cross-validation training folds were aggregated, and the final cell-type probabilities were derived by averaging the softmax outputs of the model pool. This ensemble strategy was used to mitigate potential variance from individual fold initializations and to provide an overall consensus gating logic.

### Data analysis and statistics

Model performance on the independent test set was quantified using balanced accuracy and F1-scores (weighted and unweighted). The quantification fidelity of four diagnostically relevant phenotypic fractions—total blasts/nucleated cells, total lymphocytes/nucleated cells, CD20+ blasts/total blasts, and mature B cells/total lymphocytes—was assessed via mean absolute errors (MAE) and Spearman rank correlations between predicted and expert-derived values. To determine if model-to-expert deviations remained within the bounds of inter-expert variability, one-sided Wilcoxon signed-rank tests were performed to assess whether model errors significantly exceeded inter-expert disagreement, using alternating expert annotations as references and a Bonferroni-corrected 5% significance threshold to account for multiple comparisons.

## Data availability

The raw AML flow cytometry measurements and corresponding annotations used in this work were described in the original study [19] and are publicly available from Zenodo (https://zenodo.org/records/11046402). Representative ALL test measurements (specifically the best- and worst-performing model prediction cases) will be provided alongside the optimized IMPGAT model weights upon publication. The ALL clinical dataset is not publicly available due to data protection regulations. However, these data may be shared upon reasonable request, subject to institutional and ethical approvals and the execution of a formal data transfer agreement.

## Code availability

The source code and model architecture for IMPGAT, along with the optimized model weights for both the AML and ALL cohorts, will be made publicly available in a persistent repository under an MIT license upon publication. This repository will include the necessary documentation and scripts to reproduce the primary findings and model evaluations presented in this study.

## Acknowledgments

This work was supported by the European Research Council under the European Union’s Horizon 2020 Research and Innovation Programme (grant agreements 866411, 101113551, and 101213822 to C.M.). This study was also funded in part by the Deutsche Forschungsgemeinschaft (DFG, German Research Foundation)—project number 444949889 (KFO 5010 Clinical Research Unit ‘CATCH ALL’ to G.C., C.D.B., and M.Br.), the Clinician Scientist Program in Evolutionary Medicine (project number 413490537 to G.C.), and the Deutsche José Carreras Leukämie-Stiftung (grants DJCLS R 15/11 and DJCLS 06R/2019 to M.S. and M.Br.). GC gratefully acknowledges support from the Detlef Zillikens Clinician Scientist Academy of Precision Health in Schleswig-Holstein (PHSH) and the clinician Scientist Academy Kiel.

## Supplementary Material

